# A psychometric evaluation of diffusion basis spectrum imaging indicates white matter inflammation in depression

**DOI:** 10.64898/2026.03.25.26349306

**Authors:** Luisa Kaluza, Anne Kühnel, Ekaterina Kuskova, Kim Studener, Denise Rommel, Jana Lieberz, Nils B. Kroemer

**Affiliations:** Section of Medical Psychology, Department of Psychiatry and Psychotherapy, University Hospital Bonn, University of Bonn, Bonn, Germany; Department of Psychiatry and Psychotherapy, Tübingen Center for Mental Health, University of Tübingen, Tübingen, Germany; German Center for Mental Health (DZPG), partner site Tübingen; German Center for Diabetes Research (DZD), Neuherberg, Germany; Department of Neuromuscular Diseases, Centre for Neurology, University Hospital Bonn, University of Bonn, Bonn, Germany; Department of Psychiatry and Psychotherapy, University Hospital Bonn, University of Bonn, Bonn, Germany

## Abstract

An inflammatory subtype of major depressive disorder (MDD) is associated with treatment resistance pointing to an unmet need for adjunctive treatments. To evaluate treatment-related changes in brain inflammation, diffusion basis spectrum imaging (DBSI) is a promising non-radiation-based technique for longitudinal designs which has been verified with histopathology. However, its use as an endpoint in clinical trials is dependent on its individual-level reliability to robustly track changes. Here, we evaluated two DBSI runs acquired in 94 participants (including 43 participants with MDD) on the same day about 1.5 h apart to assess short-term test–retest reliability. Fiber fraction (reflecting axonal/dendrite density) and hindered fraction (reflecting edema) showed moderate to high test–retest reliability in both gray and white matter regions, whereas restricted fraction (reflecting cellularity) showed lower values in gray and white matter. Group-level reliability was similar in participants with MDD, except for lower reliability of hindered fraction in gray matter. Re-identification rates of individual brain maps were higher using voxel-level white matter signatures compared to gray matter regions of interest (ROIs) (*p*<.001). Crucially, participants with MDD showed reduced fiber fraction (*t*_max_=4.68, *k*=38) and elevated hindered fraction (*t*_max_=4.74, *k*=32) in the cingulate bundle, consistent with increased white matter inflammation, while gray matter ROI-based classification failed to identify cases. We conclude that DBSI is a promising technique to track inflammatory signatures in MDD, particularly in white matter tracts. Since several frontal and subcortical gray matter ROIs showed insufficient reliability, their assessment would require multiple DBSI runs to provide robust estimates.

## 1. Introduction

Major depressive disorder (MDD) is thought to encompass multiple subtypes with partially distinct etiologies and treatment responses (Akil et al., 2018; Liang et al., 2025; Nguyen et al., 2022; Wang et al., 2025). An inflammatory subtype has recently gained traction due to its resistance to conventional treatments requiring new adjunctive options (He et al., 2026; Treadway et al., 2024) and its alleged association with obesity (Chan et al., 2019). Induction of depressive-like behavior and alterations in dopaminergic or reward-related signaling have been demonstrated following inflammatory stimuli in animal (Felger et al., 2013, 2007; Felger and Treadway, 2017) and human studies (Capuron et al., 2012, 2007; Eisenberger et al., 2010; Felger and Treadway, 2017). Accordingly, L-DOPA interventions increased functional connectivity within reward circuits in patients with high baseline markers of peripheral inflammation (Bekhbat et al., 2024, 2022). In addition, higher levels of inflammatory markers in patients with MDD have been linked to altered signaling during reward anticipation (Burrows et al., 2021) and reduced functional connectivity within the cortico-striatal reward circuit associated with symptoms, such as anhedonia and psychomotor slowing (Felger et al., 2016). To conclude, emerging evidence has emphasized the need to measure inflammation in patients with MDD. Whereas peripheral inflammatory markers are often used for inclusion in clinical trials (Giollabhui et al., 2025), repeated quantification of central inflammation may complement such approaches to advance treatment stratification or evaluate their effects over time.

In humans, central inflammation is usually assessed using neuroimaging techniques (Gritti et al., 2021). Although central inflammation could be demonstrated in MDD across a variety of studies using positron emission tomography (for review see: Meyer et al., 2020), it is not feasible for repeated measurements due to high costs, cumulative radiation exposure (especially in combination with computed tomography), and limited availability (Berger et al., 2003; Griffeth, 2005). Magnetic resonance imaging (MRI), including diffusion-based approaches, offers a radiation-free alternative to investigate central inflammation (Oestreich and O’Sullivan, 2022). To study microstructure white matter changes, diffusion tensor imaging (DTI) has been extensively evaluated in patients with MDD (Guo et al., 2024; Liao et al., 2013). However, conventional DTI lacks specificity to identify pathologies (Alexander et al., 2007). Diffusion kurtosis imaging extends DTI by accounting for deviations from a Gaussian distribution, thereby taking biological complexity into account (Lu et al., 2006). Moreover, free water mapping may help extend the applicability of DTI to tissues containing cerebrospinal fluid or edema, providing additional quantitative diffusion-based measurements (Pasternak et al., 2009). These measures point to neuroinflammation in depression (Langhein et al., 2022; Oestreich et al., 2020). In addition, neurite orientation dispersion and density imaging may further improve the specificity of microstructural changes in white matter by adding neurite density and orientation dispersion measures (Zhang et al., 2012) with demonstrated changes in MDD compared to healthy control participants (HCP) (Khalife et al., 2025; Ota et al., 2018; Takahashi et al., 2025). In contrast to other diffusion-based protocols, diffusion basis spectrum imaging (DBSI) assesses additional proxies of neuroinflammation, such as cellularity (Wang et al., 2011). Rodent studies have shown associations between histology measures (e.g., cell counts) and proxies of cellularity determined with DBSI (Wang et al., 2011). Likewise, DBSI-related proxies of central inflammation in gray and white matter have been associated with obesity (Dolatshahi et al., 2025; Samara et al., 2021, 2019). Some studies have used DBSI in patients with MDD or in association with depressive symptoms to identify, for example, increased restricted fraction in the amygdala (Kaszás et al., 2025; Zhang et al., 2023). Accordingly, DBSI may provide a promising technique to repeatedly assess central inflammation, but its psychometric quality has not been formally tested in patients with MDD.

Reliability and statistical power are practically relevant and interconnected factors guiding the design of an informative trial (Liem et al., 2015). Although statistical power can be estimated from an anticipated (latent) effect size, evaluating reliability requires sufficient data, ideally from test-retest settings. Reliability influences the *observed* effect size by capturing an attenuation of the anticipated effect size due to measurement noise. This affects the required sample size given the reliability of a given measure. Reliability assessments have been conducted for diffusion kurtosis imaging and DTI metrics, generally demonstrating good reliability, although with region-specific variability and differences among diffusion metrics (Luque Laguna et al., 2020; Wu et al., 2022). However, the test-retest reliability of DBSI has not been quantified. To address this critical gap, we acquired two identical DBSI runs approximately 1.5 h apart in 94 participants (including 43 participants with MDD). Our results demonstrate overall moderate to high test-retest reliability in white matter and many gray matter regions of interest (ROIs) with DBSI fiber fraction (i.e., a proxy for dendritic and axonal density) showing the best reliability. Likewise, DBSI fiber fraction achieved the highest re-identification rates at the single-subject level. Crucially, participants with MDD showed reduced fiber fraction and elevated hindered fraction in the cingulate bundle, while gray matter ROIs did not robustly classify cases vs. controls. Hence, our study demonstrates that DBSI provides reliable voxel-level information on proxies of white matter inflammation, and mostly reliable estimates in gray matter ROIs, although averaging may be necessary for frontal or subcortical target regions.

## 2. Methods

### 2.1 Participants

For the reported analyses, we included DBSI data of an ongoing larger study investigating the acute and medium-term effects of transcutaneous vagus nerve stimulation (tVNS) in patients with MDD (pre-registered on clinicaltrials.gov, ID: NCT06389175). At the time of the data freeze for DBSI in February 2026, 100 participants had completed both neuroimaging sessions, with complete DBSI data available in 94 participants (DBSI was not acquired due to technical problems in the other participants). The sample included 51 HCP (29 women, *M_age_*=31.6 years ±9.9) without a lifetime history of depression and 43 participants currently fulfilling the criteria for MDD (20 women, *M_age_*=32.4 years ±8.4, Table 1). The reported analyses and results were not included in the preregistration. The study received approval from the ethics committee of the Faculty of Medicine at the University of Bonn (07/28/2022, 232/22) in accordance with the Declaration of Helsinki, and each participant provided written informed consent prior to participation. The privacy rights of human subjects have been observed.

**Table 1.** Participant characteristics.

|  | **HCP, *n* = 51** | **MDD, *n* = 43** | ***p*-value** |
| --- | --- | --- | --- |
| **Age [years]** | 31.6 ± 9.9<br>[18 – 55] | 32.4 ± 8.4<br>[18 – 54] | 0.68<sup>1</sup> |
| **Sex** |  |  | 0.43<sup>2</sup> |
| Male, *n* (%) | 22 (43.1%) | 23 (53.5%) |  |
| Female, *n* (%) | 29 (56.9%) | 20 (46.5%) |  |
| **BMI [kg/m<sup>2</sup>]** | 23.5 ± 2.8<br>[18.5 – 29.6] | 24.1 ± 2.9<br>[18.2 – 30.5] | 0.32<sup>1</sup> |
| **Stimulation between DBSI runs** |  |  | >0.99<sup>2</sup> |
| Sham, *n* (%) | 24 (47.1 %) | 20 (46.5 %) |  |
| tVNS, *n* (%) | 27 (52.9 %) | 23 (53.5 %) |  |
| **BDI<sup>a</sup>** | 5.9 ± 5.9<br>[0 – 26] | 27.2 ± 9.7<br>[8 – 44] | <0.001<sup>1</sup> |
| Minimal, *n* (%) | 45 (88.2 %) | 2 (4.7 %) |  |
| Mild, *n* (%) | 4 (7.8 %) | 5 (11.6 %) |  |
| Moderate, *n* (%) | 2 (3.9 %) | 15 (34.9 %) |  |
| Severe, *n* (%) | 0 (0.0 %) | 18 (41.9 %) |  |
| **Anti-depressive medication** |  |  |  |
| SSRI, *n* (%) | 0 (0.0 %) | 8 (18.6 %) | 0.001<sup>3</sup> |
| Other, *n* (%) | 0 (0.0 %) | 8 (18.6 %) | 0.001<sup>3</sup> |
| None, *n* (%) | 51 (100 %) | 27 (62.8 %) | <0.001<sup>3</sup> |
| **Comorbidities** |  |  |  |
| Anxiety-related, *n* (%) | 11 (21.6 %) | 24 (55.8 %) | 0.001<sup>3</sup> |
| Psychosis-related, *n* (%) | 0 (0.0 %) | 1 (2.3 %) | 0.46<sup>3</sup> |
| Substance abuse (mild), *n* (%) | 2 (3.9 %) | 0 (0.0 %) | 0.50<sup>3</sup> |
| Trauma-related, *n* (%) | 0 (0.0 %) | 4 (9.3 %) | 0.04<sup>3</sup> |
| ADHD, *n* (%) | 0 (0.0 %) | 11 (25.6 %) | <0.001<sup>3</sup> |

**Note:** Data are means ± SD [range] if not indicated otherwise. *Abbreviations:* ADHD = attention deficit hyperactivity disorder, BDI = Beck Depression Inventory, BMI = body mass index, DBSI = diffusion basis spectrum imaging, HCP = healthy control participants, MDD = major depressive disorder, SSRI = selective serotonin reuptake inhibitor, tVNS = transcutaneous vagus nerve stimulation. <sup>a</sup>BDI data of *n*=3 were missing. <sup>1</sup>Student's two-sample *t*-test; <sup>2</sup>Chi-squared test; <sup>3</sup>Fisher's exact test.

### 2.2 Experimental procedure

DBSI data was acquired during the second neuroimaging session (Fig. 1). For both neuroimaging sessions, participants came in after an overnight fast. Then, anthropometric measurements (weight and height) and a blood sample were taken. Afterwards, participants performed a food cue reactivity task (Müller et al., 2022). During the second neuroimaging session, we acquired two DBSI scans: at the beginning of the MRI assessment (instead of the structural T1 scan acquired during the first session) and at the end. In between, functional MRI was measured during resting state (20 min, Vanderwal et al., 2015), tasks (∼45 min), and another resting state (10 min), with ∼65 min of concurrent tVNS or sham stimulation (see SI). Participants answered state questions (e.g., happy and sad) on a visual analog scale (Ferstl et al., 2022) three times throughout the session. There was a washout period of about one week (*M*=7.62 d ±5.67, range 2–35) between neuroimaging sessions.

**Figure 1:**
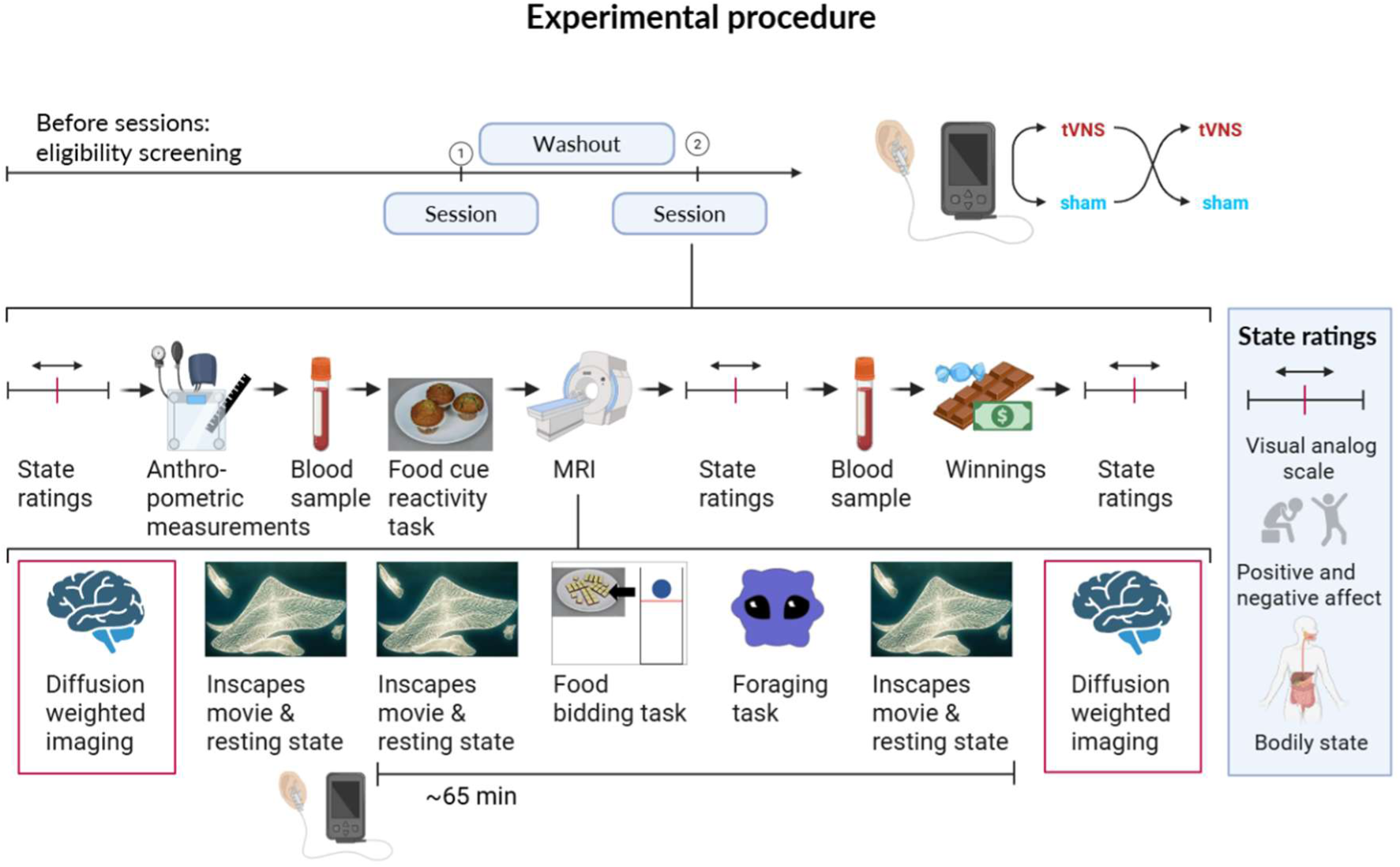
Experimental procedure. Participants came in after an overnight fast. State ratings, anthropometric measurements, blood sampling, and a food cue reactivity task were performed before the MRI assessment. The same diffusion weighted imaging sequence was acquired twice during the second neuroimaging session while functional MRI was collected in between. After the first diffusion basis spectrum imaging run, participants received ∼65 min of stimulation with either transcutaneous vagus nerve stimulation (tVNS) or sham.

### 2.3 MRI data acquisition and preprocessing

MRI data were acquired with a 3 Tesla MRI scanner (Siemens Magnetom Trio) using a 32-channel head coil. For each participant, we acquired the same diffusion weighted imaging (DWI) sequence twice: planar AP DWI images (voxel dimensions: 2 x 2 x 2 mm, TE = 95 ms, TR = 10,000 ms, transverse orientation, flip angle = 90°, 26 directions, b-values ranging from 0 to 1400 s/mm^2^) and two non-diffusion images (b-value = 0) in AP and PA direction, respectively. In addition, one high-resolution T1-weighted (T1w) anatomical image (208 sagittal slices, flip angle = 9°, matrix size = 256 x 256 and voxel size = 0.8 x 0.8 x 0.8 mm³) was acquired during the first neuroimaging session for each participant.

DWI images of the runs were preprocessed separately with FSL (Jenkinson et al., 2012). First, we combined the two non-diffusion images (AP and PA direction). Afterwards, distortions were corrected using `topup` (Andersson et al., 2003), and `eddy_correct` was applied to correct for eddy currents and head motions. We then calculated DBSI maps (Wang et al., 2011) that capture three indices of central inflammation. First, fiber fraction measures the anisotropic diffusion component which represents the water molecules inside and outside but adjacent to axon fibers and estimates axonal or dendrite density. Second, cellularity is assessed by measuring the isotropic diffusion of water molecules inside cells or in sub-cellular structures and thus appears to be highly restricted (restricted fraction). Third, hindered fraction assesses isotropic diffusion of water molecules that are less restricted and have been associated with vasogenic edema (Cross and Song, 2017; Wang et al., 2011). In line with previous studies (Samara et al., 2021), the isotropic diffusion spectrum *f(*D) with D≤0.3 μm^2^/ms reflects the restricted fraction and D=0.3–3.0 μm^2^/ms hindered fraction.

The T1w images were preprocessed using fMRIprep 23.2.3 (Esteban et al., 2019; for full output, see SI), including correction for intensity non-uniformity, volume-based spatial normalization to standard space (MNI152NLin2009cAsym), and brain-extraction. To normalize DBSI maps to MNI space, diffusion-derived b0 images were nonlinearly registered to the individual T1-weighted anatomical image using `antsRegistrationSyNQuick.sh`, with the skull-stripped T1-weighted image from fMRIprep as reference. The resulting transformations were combined with the precomputed T1w-to-MNI152NLin2009cAsym nonlinear warp and applied with `antsApplyTransforms`.

### 2.4 Data analysis

#### 2.4.1 Reliability of DBSI

To estimate the test-retest reliability of the three DBSI parameters (fiber fraction, hindered fraction, and restricted fraction), we first calculated the Spearman correlation between the estimated parameters from both runs. For gray matter, we calculated the reliability for each region after averaging across voxels within each region of the extended Harvard-Oxford Brain Atlas, including subcortical regions of the Reinforcement Learning Atlas and the AAL cerebellum (Desikan et al., 2006; Pauli et al., 2018; Teckentrup et al., 2021). For white matter, we calculated the Spearman correlation in each voxel within a white matter mask (voxels with SPM12 tissue *p*>.50). As in previous work (Fröhner et al., 2019), we interpret test-retest correlations as low (≤0.35), moderate (0.36-0.67), or high (>0.67) according to Taylor (1990). To compare the reliability between groups, we used the Kolmogorov-Smirnov test to compare the distributions across ROIs.

Next, to determine whether the individual information contained in the DBSI maps across runs is sufficient to re-identify participants, we used a ‘fingerprinting’ approach (Finn et al., 2015; Fröhner et al., 2019). Briefly, the spatial pattern of an individual brain map of a run is correlated with the brain map of the same individual of the second run, as well as with the brain maps of all other participants of the second run. This gives a correlation matrix, in which the diagonal values correspond to the correlation of a spatial pattern within an individual (i.e., self-correlation) and the off-diagonal values capture the similarity with brain maps of other individuals. Re-identification rate refers to the percentage of individuals for whom the diagonal value (i.e., self-correlation) is higher than any off-diagonal value in the same column (i.e., other correlation). We used this approach based on separate patterns of the gray matter atlas ROIs or white matter voxels.

#### 2.4.2 Interindividual differences in central inflammation

To test voxel-wise individual differences in any of the DBSI parameters in white matter, we used full factorial models in SPM12 (Wellcome Department of Imaging Neuroscience, University College London, London, UK Friston et al., 2007). The models included two factors: Run (before and after stimulation, dependent measurements) and MDD (HCP vs. MDD, independent measurements), as well as covariates (all grand mean centered) indicating stimulation (tVNS vs. sham), age, BMI, and sex. The comparisons of interest were MDD vs. HCP and the correlations with BMI and age across groups.

To test the predictive power of DBSI parameters based on gray matter ROIs, we used elastic net prediction models (*lasso*, preset alpha=.5, Matlab2024a) and nested 10-fold cross-validation. As in previous studies (Kühnel et al., 2025, 2023, 2022), we used an elastic net which performs well with a moderate number of correlated features (Jollans et al., 2019). To reduce the number of features, any bilateral region was averaged across the left and right hemispheres, leading to 82 features. First, we predicted individual age as a benchmark, since age is highly associated with changes in brain morphology and inflammation (Bennett et al., 2009; Jurcau et al., 2024). We then predicted both BMI and MDD (vs. HCP, with a generalized elastic net model) using the same set of predictors. Statistical significance was determined using permutation tests with 1,000 iterations.

#### 2.4.3 Statistical threshold and software

We used a significance threshold of *p*<.05 (two-tailed) for all main outcomes. White matter group-level analyses were tested using a voxel threshold of *p*<.001 and cluster-level family-wise error (FWE) correction (*p*_FWE_<.05). Additional statistical analyses and data visualization were completed with R v4.4.2 (R Core Team, 2024) using the following packages: beeswarm (Eklund and Trimble, 2021), lmerTest (Kuznetsova et al., 2017), ggplot2 (Wickham, 2016), cowplot (Wilke, 2024), and viridis (Garnier et al., 2024).

## 3. Results

### 3.1 Higher test-retest reliability of fiber and hindered fraction

First, we determined the test-retest reliability of DBSI metrics across two scans collected within 1.5 h on the same day. To estimate the stability of the rank order, we calculated Spearman correlations for gray matter ROIs and white matter voxels (Fig. 2). For white matter voxels (i.e., the conventional focus of DWI), we mostly observed moderate to high reliability for fiber fraction (15.5% with high and 60.6% with moderate reliability) and hindered fraction (3.5% with high and 63.1% with moderate reliability), particularly in voxels closer to gray matter and in the brainstem. In contrast, restricted fraction predominantly showed low reliability (88% of white matter voxels). For gray matter ROIs, we observed a similar pattern with moderate to high reliabilities for fiber fraction and hindered fraction (>95% of ROIs with moderate reliability or better). In contrast, >25% of ROIs showed low reliability for restricted fraction. Across DBSI parameters, some subcortical gray matter ROIs that are implicated in MDD or obesity, such as the nucleus accumbens (NAcc), the ventral pallidum, and the ventral tegmental area (VTA) showed insufficiently low reliability, which was much lower compared to cortical regions including the insula, superior parietal lobule, and sensorimotor cortex.

**Figure 2:**
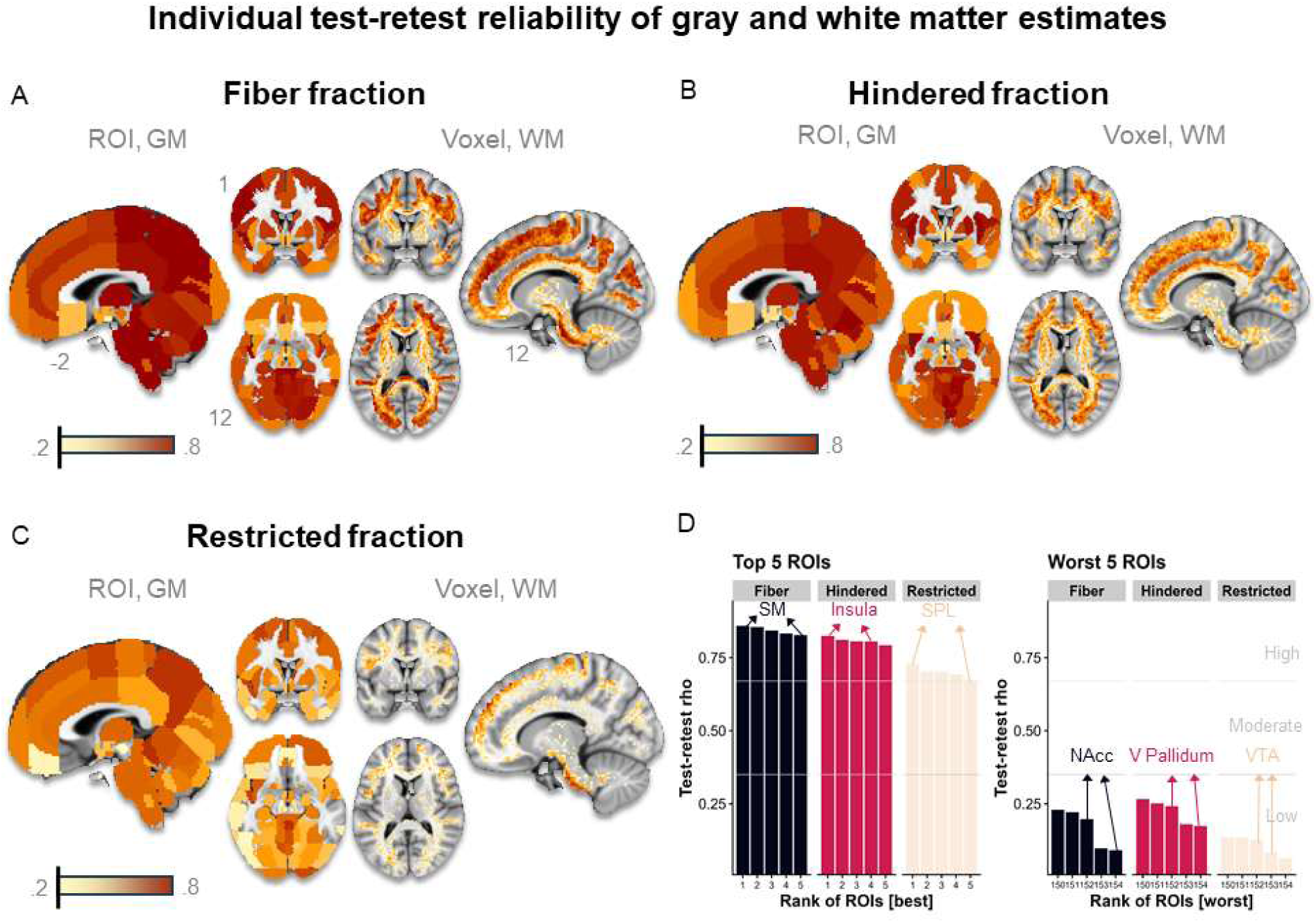
Test-retest reliability of gray and white matter estimates. A: Fiber fraction demonstrated moderate to high reliability for both gray matter (GM; >95% of ROIs with moderate reliability or better) and white matter (WM; 15.5% of white matter voxels with high and 60.6% with moderate reliability). B: Hindered fraction also demonstrated moderate to high reliability for gray matter (>95% of ROIs with moderate reliability or better) and white matter (3.5% of white matter voxels with high and 63.1% with moderate reliability). C: Restricted fraction showed the lowest values with low reliability in >25% of gray matter ROIs and 88% of white matter voxels. D: Subcortical gray matter regions, such as the nucleus accumbens (NAcc), ventral pallidum (V Pallidum), and the ventral tegmental area (VTA) showed lower reliability estimates as compared to cortical gray matter regions. The color bar indicates rho. ROI = region of interest, SM = sensorimotor cortex, SPL = superior parietal lobule.

Second, we tested for differences in reliability between patients with MDD vs. HCP and found no consistent differences for white matter voxels (i.e., distributions of differences were mostly centered around 0). However, the reliability of gray matter ROIs was significantly lower in participants with MDD for hindered fraction (*D*(51,43)=0.25, *p*<.005, Fig. S1) whereas there were no differences in other DBSI metrics.

### 3.2 Re-identification of individuals is higher using white vs. gray matter

To determine whether DBSI metrics can be used to re-identify participants, we used a fingerprinting approach and calculated within- and between-subject correlations of the two runs. For white matter voxel maps, re-identification rates (i.e., the percentage of participants for which the correlation with themselves was higher than the correlation with any other participant) were very high (ID_Rate_>93%) for all three measures (Fig. 3B, D). The self-correlation values indicated high test-retest correlations for fiber fraction (*Mr*_self_=.74) and hindered fraction (*Mr*_self_=.70), but they were low for restricted fraction (*Mr*_self_=.30). The latter results indicate that although individuals can be re-identified, it is insufficiently accurate for tracking changes over time. For gray matter ROIs, re-identification rates ranged between 73.0% for fiber fraction, 71.2% for hindered fraction, and 45.7% for restricted fraction, all significantly lower than for white matter voxels (all *p*s<.001, Fig. 3A, C). The self-correlation values were higher compared to white matter voxels with all metrics showing high average correlations (fiber fraction: *Mr*_self_=.88, hindered fraction: *Mr*_self_=.92, restricted fraction: *Mr*_self_=.68). However, off-diagonal correlations were also much higher on average, indicating that gray matter ROIs contained less unique information compared to white matter voxels. Overall, fiber fraction and hindered fraction showed the highest re-identification rates for both white matter voxels and gray matter ROIs together with high test-retest correlations indicating suitability to detect interindividual differences and track changes over time.

**Figure 3:**
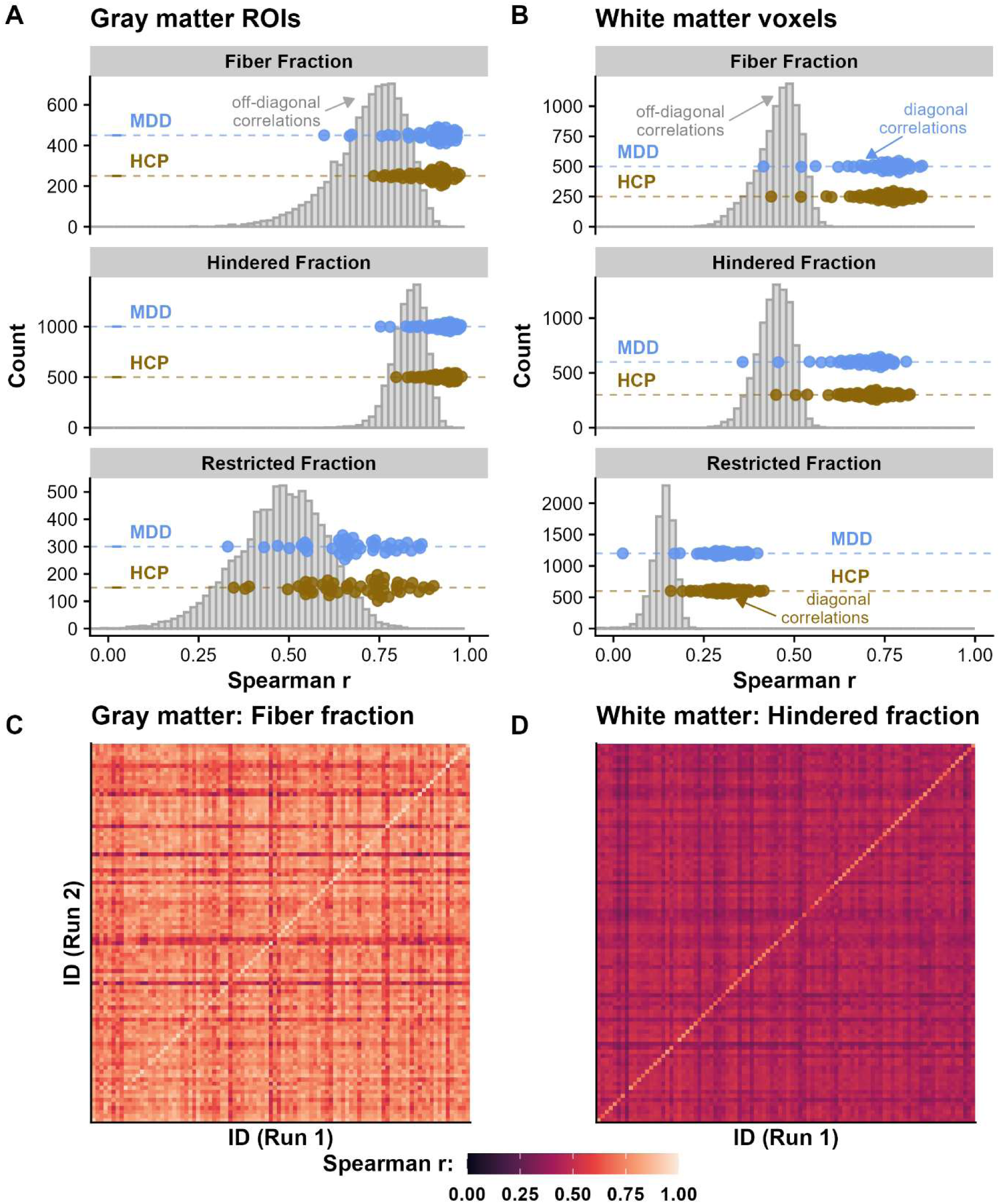
Re-identification rates of individual brain maps are higher for white compared to gray matter. Re-identification rates were estimated by calculating correlations between individual diffusion basis spectrum imaging (DBSI) maps. Diagonal values (blue: major depressive disorder, golden: healthy control participants) reflect correlation of a spatial pattern within an individual, and off-diagonal values (gray histograms) reflect the correlation with other individuals. A: For gray matter regions of interest (ROIs), fiber fraction (73.0%) and hindered fraction (71.2%) showed high re-identification rates, whereas restricted fraction showed a lower re-identification rate (45.7%). All were higher than chance level (1%). B: Re-identification rates were significantly higher for white matter voxels (>93%, ps<.001). However, restricted fraction showed low self-correlations (*Mr*_self_=.30), indicating low precision. C: Rank correlation matrix of fiber fraction patterns across gray matter ROIs between Run 1 and Run 2 across participants. D: Rank correlation matrix of hindered fraction of white matter voxel maps from Run 1 and Run 2 across participants.

### 3.3 Higher inflammation in the cingulum bundle in participants with MDD

Next, we assessed whether participants with MDD vs. HCP showed changes in central inflammation in white matter voxels using factorial models in SPM. Participants with MDD showed reduced fiber fraction (*t*_max_=4.68, *k*=38, *p*_FWE_<.001) and elevated hindered fraction (*t*_max_=4.74, *k*=32, *p*_FWE_<.001) in the right cingulate bundle, indicating a loss of axonal density and an increase in tissue edema (Fig. 4). In addition, fiber fraction was reduced in white matter clusters in the visual cortex (*t*_max_=4.90, *k*=29, *p*_FWE_=.001) and the supplementary motor area (*t*_max_=4.29, *k*=26, *p*_FWE_=.002). Hindered fraction was also increased in white matter cluster in the visual cortex (*t*_max_=4.78, *k*=27, *p*_FWE_<.001). Since these clusters were in regions with high voxel-level test-retest reliability, those interindividual differences are supported by reliable inter-individual estimates.

**Figure 4:**
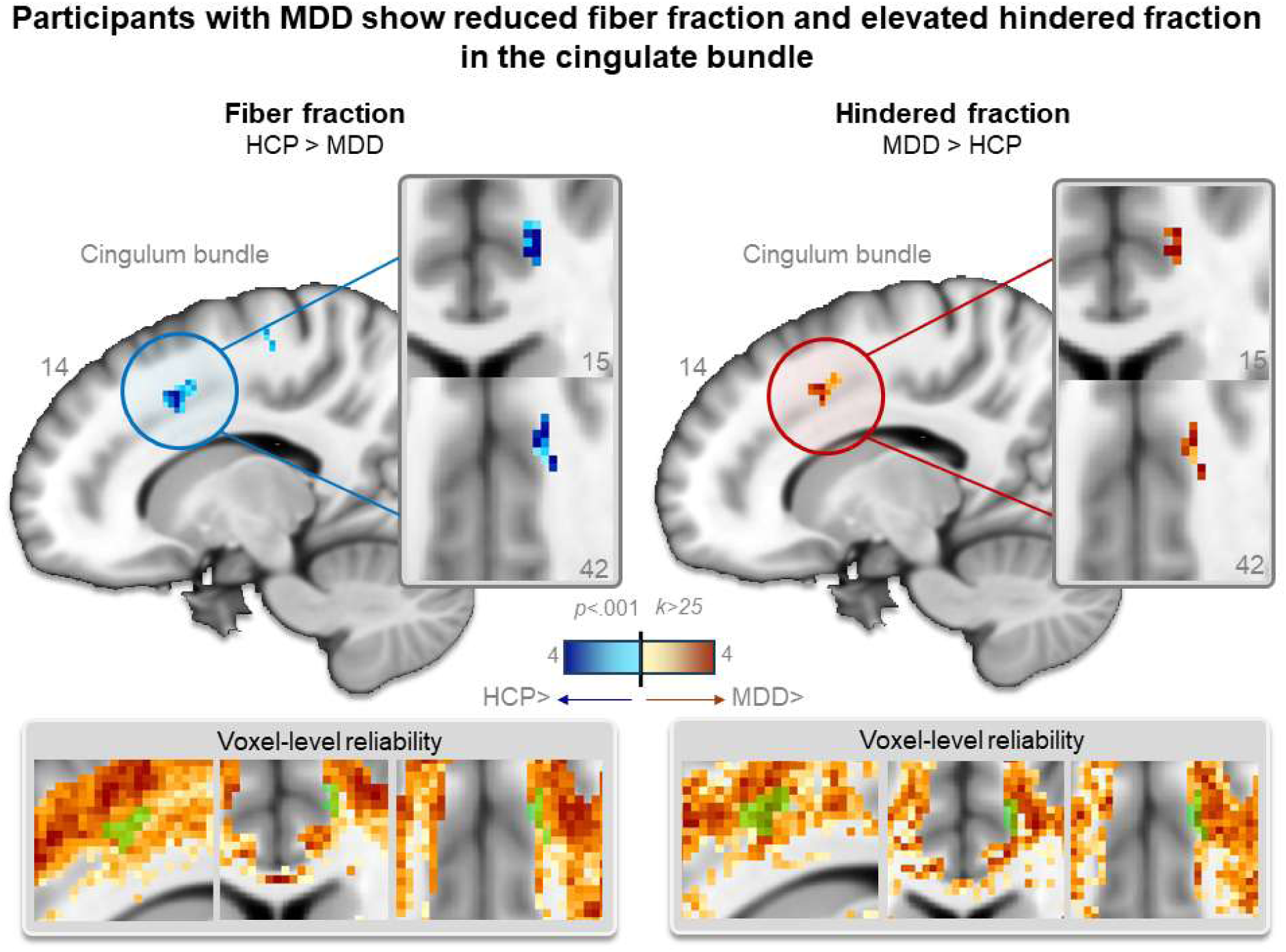
Participants with major depressive disorder (MDD) show lower fiber fraction and higher hindered fraction in the cingulum bundle. We observed differences in the cingulum bundle with decreased fiber fraction (*t*_max_=4.68, *k*=38, *p*_FWE_<.001) indicating reduced axonal integrity (left) and increased hindered fraction (*t*_max_=4.74, *k*=32, *p*_FWE_<.001), suggesting edema (right) in participants with MDD vs. healthy control participants (HCP). These changes indicate increased white matter inflammation in participants with MDD. In voxels showing differences between HCP and MDD groups, the reliability of fiber fraction and hindered fraction was high (lower insets).

### 3.4 Gray matter ROIs patterns of inflammation predict age, not BMI

To determine the predictive accuracy of central inflammation in gray matter ROIs, we used individual patterns as features to predict age (as an established benchmark), BMI, and MDD (vs. HCP). Features from all three DBSI measures successfully predicted age above chance levels (fiber fraction: *R²*=.22, *p*_perm_<.001, hindered fraction: *R²*=.36, *p*_perm_<.001, restricted fraction: *R²*=.26, *p*_perm_<.001) explaining between 22% and 36% of the variance (Fig. 5A-B). In contrast, neither BMI (*p*s_perm_>.29, Fig. 5C) nor MDD status (vs. HCP, *p*s_perm_>.63, Fig. 5D) was predicted significantly above chance levels by any of the DBSI metrics. Taken together, the results suggest that DBSI gray matter ROI metrics contain sufficient individual information to predict individual differences, although the prediction of BMI or disorder status solely based on gray matter ROIs may require larger samples.

**Figure 5:**
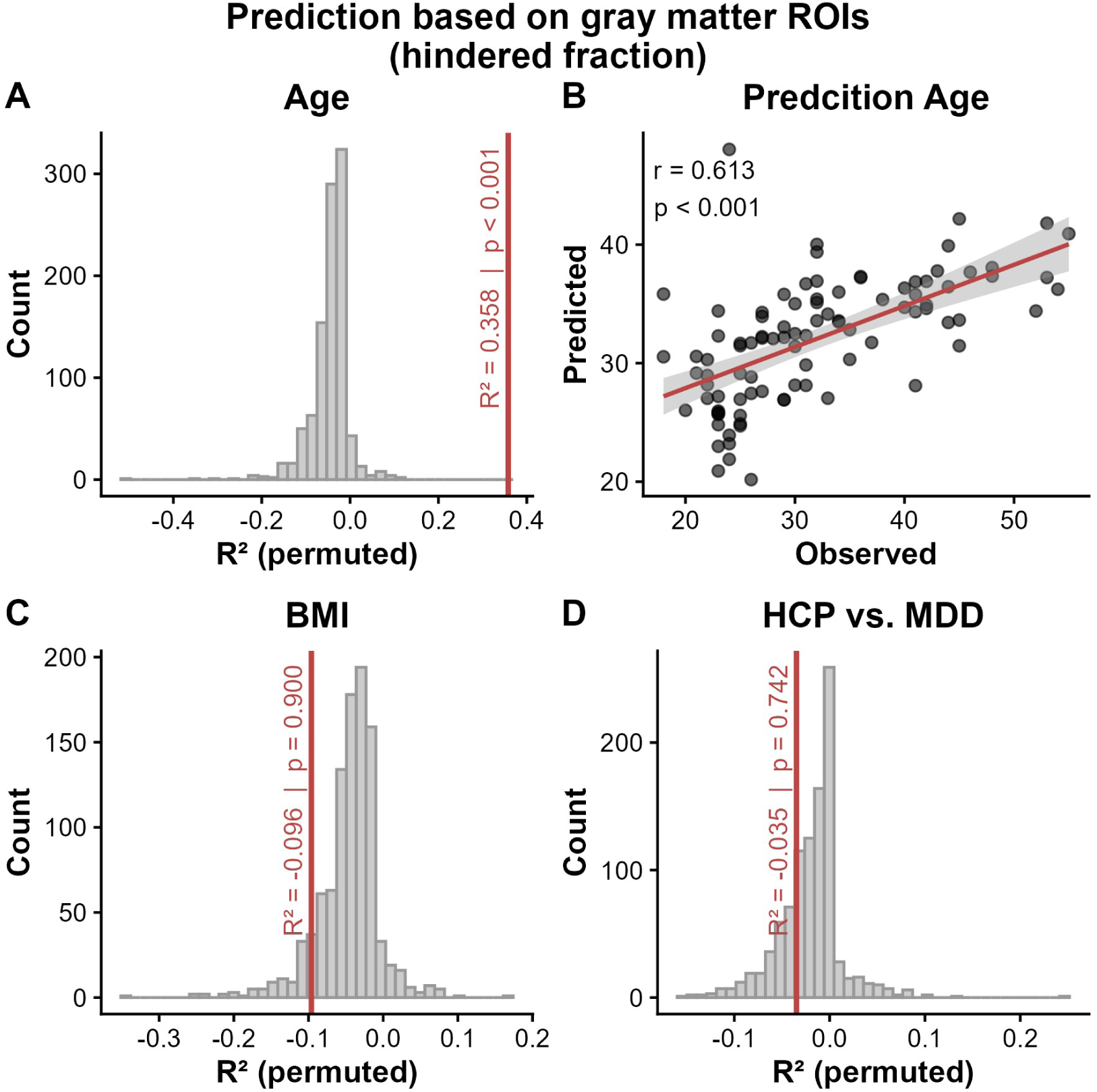
Diffusion basis spectrum imaging (DBSI) metrics predict age. A: Gray matter region of interest (ROI) features of hindered fraction successfully predicted age above chance level (*R²*=.36, *p*_perm_<.001). B: Predicted values of age were highly correlated with the observed age (*r*=.61, *p*<.001). C: Gray matter ROI features of hindered fraction failed to predict BMI beyond chance level (*p*s_perm_>.29). D: Gray matter ROI features of hindered fraction did not predict whether a participant had a major depressive disorder (MDD) or was a healthy control participant (HCP) beyond chance level (*p*s_perm_>.63). In all panels, the red line indicates the observed predictive performance of the model, and the histogram depicts the distribution of the permutations. Results for other DBSI metrics were comparable; hindered fraction is shown here as a representative example.

## 4. Discussion

MDD is a highly heterogeneous condition, and concurrent inflammation is associated with treatment resistance to antidepressants. DBSI promises to measure several proxies of neuroinflammation across repeated measurements without radiation or contrast agents, which could complement the measurement of peripheral inflammation in future trials to improve the evaluation of treatments targeting inflammation. However, the test-retest reliability of DBSI has not been evaluated yet, although the robust estimation of individual brain maps is a prerequisite for its validity, specifically in randomized controlled trials (RCTs). To address this gap, we evaluated short-term test–retest reliability and re-identification rates of individual brain maps across two runs in participants with MDD and HCP. Re-identification rates of individual brain maps were high for fiber fraction and hindered fraction, particularly for voxel-level white matter signatures. As expected, participants with MDD showed lower fiber fraction and higher hindered fraction in the cingulate bundle, consistent with inflammation in white matter tracts. In contrast, using gray matter ROIs alone, we could not robustly classify patients vs. controls, whereas age was predicted with good accuracy with all DBSI estimates. We conclude that DBSI provides reliable and unique information on individual proxies of white matter inflammation even at the voxel level, whereas averaging may be necessary to boost reliability in gray matter ROIs, particularly in frontal and subcortical regions.

In line with previous DWI studies, we found that participants with MDD had reduced fiber fraction and increased hindered fraction in the cingulate bundle. Reduced fractional anisotropy indicating white matter microstructure changes in the cingulum has been repeatedly observed in depression (Barch et al., 2022; de Diego-Adeliño et al., 2014; van Velzen et al., 2020), and changes in fractional anisotropy in the cingulum over time have been associated with changes in depressive symptoms (Doolin et al., 2019). Crucially, cingular white matter tract integrity is reduced with higher levels of peripheral inflammatory markers in MDD (Thomas et al., 2022). Moreover, microstructure changes in the cingulum have been associated with treatment resistance, chronic disease status (de Diego-Adeliño et al., 2014), fatigue, and anhedonia (Coloigner et al., 2019; Keedwell et al., 2012; Pardini et al., 2015). In turn, anhedonia has been associated with inflammation, changes in dopaminergic signaling, and treatment resistance (Felger and Treadway, 2017; Varela et al., 2025). Our study extends previous findings by showing increased hindered fraction reflecting edema (Cross and Song, 2017; Samara et al., 2021; Wang et al., 2011) in the cingulate bundle. To summarize, our results suggest that white matter changes in the cingulum bundle are driven by edema, which is reflected in increased hindered fraction with DBSI.

Despite significant differences between participants with MDD and HCPs in the cingulum bundle, we did not observe robust differences in gray matter ROIs. Overall, we observed moderate to high test–retest reliability for fiber fraction and hindered fraction and consistently lower values for restricted fraction in both gray and white matter regions. Previous studies have shown that DWI measures show different psychometric properties in white and gray matter, with lower values typically observed in gray matter (Mito et al., 2026; Seider et al., 2022). Several subcortical gray matter ROIs produced insufficiently low reliability, including key regions associated with reward and motivation and symptoms of anhedonia, such as NAcc and VTA (Liu et al., 2021; Stelzner et al., 2025; Wacker et al., 2009). Likewise, re-identification rates were higher for white matter voxels compared to gray matter ROIs, and higher compared to typical fMRI measures, thereby enabling longitudinal assessments with great precision at the individual level (Fröhner et al., 2019). Small within-subject vs. between-subject variability has been considered a prerequisite to capture interindividual differences and track within-subject changes over time with DWI (Cole et al., 2014; Luque Laguna et al., 2020) and our re-identification rates illustrate the potential of DBSI. Thus, we conclude that DBSI-derived measures from a single run provide reliable measures in white matter voxels and most gray matter ROIs for fiber fraction and hindered fraction, while averaging multiple DBSI runs might be necessary to provide robust estimates for restricted fraction or several gray matter ROIs.

Our first study on the psychometric quality of DBSI has strengths and limitations that should be considered in future work. First, previous studies have shown associations of peripheral inflammation with white matter structural changes, for example, in integrity (Thomas et al., 2022). However, measures of peripheral inflammation were not available in this sample and future research should examine whether DBSI-related alterations in MDD are linked to peripheral inflammatory markers. Second, participants received an acute stimulation between both DBSI runs. We conducted control analyses to quantify whether DBSI is sensitive to such state-related changes and found no meaningful differences between stimulation conditions (Fig. S2). Third, whereas our results provide reliability estimates that may guide future work, it is not known how strongly other sources of variance (e.g., scanner type, imaging sequence, or preprocessing procedures) affect the estimates (Thieleking et al., 2021). Fourth, our study focused on short-term test-retest reliability and comparisons between participants with MDD and HCPs as well as predictive modeling of age and BMI to assess validity. However, additional validation studies with established measures of neuroinflammation, as well as longer-term reliability assessments are needed since our measures collected on the same day likely provide an upper bound estimate.

## 5. Conclusion

Inflammation may contribute to the heterogeneous etiology of MDD, highlighting the need for refined stratification and improved longitudinal tracking in interventional RCTs. Here, we used DBSI-derived proxies of inflammatory changes to identify reduced fiber fraction and increased hindered fraction in the cingulum bundle in participants with MDD. These changes point to the potential role of white matter edema in causing the previously reported changes in cingular white matter integrity. In contrast, we observed no significant changes in gray matter ROIs, which also yielded lower re-identification rates and lower reliability estimates especially in subcortical regions compared to white matter voxels. Consequently, averaging multiple DBSI runs might be necessary to increase sensitivity in gray matter ROIs to improve the power of future studies. We conclude that DBSI measures of fiber fraction and hindered fraction provide reliable and promising insights into white matter inflammatory processes in MDD. This makes DBSI a suitable tool for future assessments of anti-inflammatory treatments in RCTs, particularly in patients with elevated peripheral inflammation.

## Data Availability

Brain maps are accessible via neurovault.

https://identifiers.org/neurovault.collection:23292

## Acknowledgement

We thank Nils Schäfer, Anna Zlobina, Jolina Selimanjin, Lisa Kirr, Antonia Varsamis, Elisabeth Freund, Malin Goddon, Noah Wegener, Johanna Fuchs, Annika Hartmann, Leyla Fox, and Christin Schwarzer for help with data acquisition. We also thank Sheng-Kwei Song for providing the Matlab code to calculate DBSI metrics. The study was supported by DFG KR 4555/9-1 and KR 4555/10-1. LK received support from DFG 493623632. AK received support from the Else Kroener Fresenius Stiftung grant 2024_EKEA.149.

## CRediT authorship contribution statement

**Luisa Kaluza:** Data Curation, Investigation, Project Administration, Writing-original draft**; Anne Kühnel:** Formal Analysis, Methodology, Validation, Visualization, Writing-original draft; **Ekaterina Kuskova:** Investigation, Writing-review & editing**; Kim Studener:** Investigation, Writing-review & editing**; Denise Rommel:** Writing-review & editing**; Jana Lieberz:** Data Curation, Investigation, Project Administration, Validation, Writing-review & editing; **Nils B. Kroemer:** Conceptualization, Formal Analysis, Funding Acquisition, Methodology, Supervision, Validation, Visualization, Writing-original draft.

## Data and code availaibility statement

Brain maps are accessible via neurovault: https://identifiers.org/neurovault.collection:23292. Analysis code is available on github: https://github.com/neuromadlab/dbsi_reliability.

