## supplements for "A psychometric evaluation of diffusion basis spectrum imaging indicates white matter inflammation in depression"

Venusberg Campus 1, 53127 Bonn, Germany

### Figures

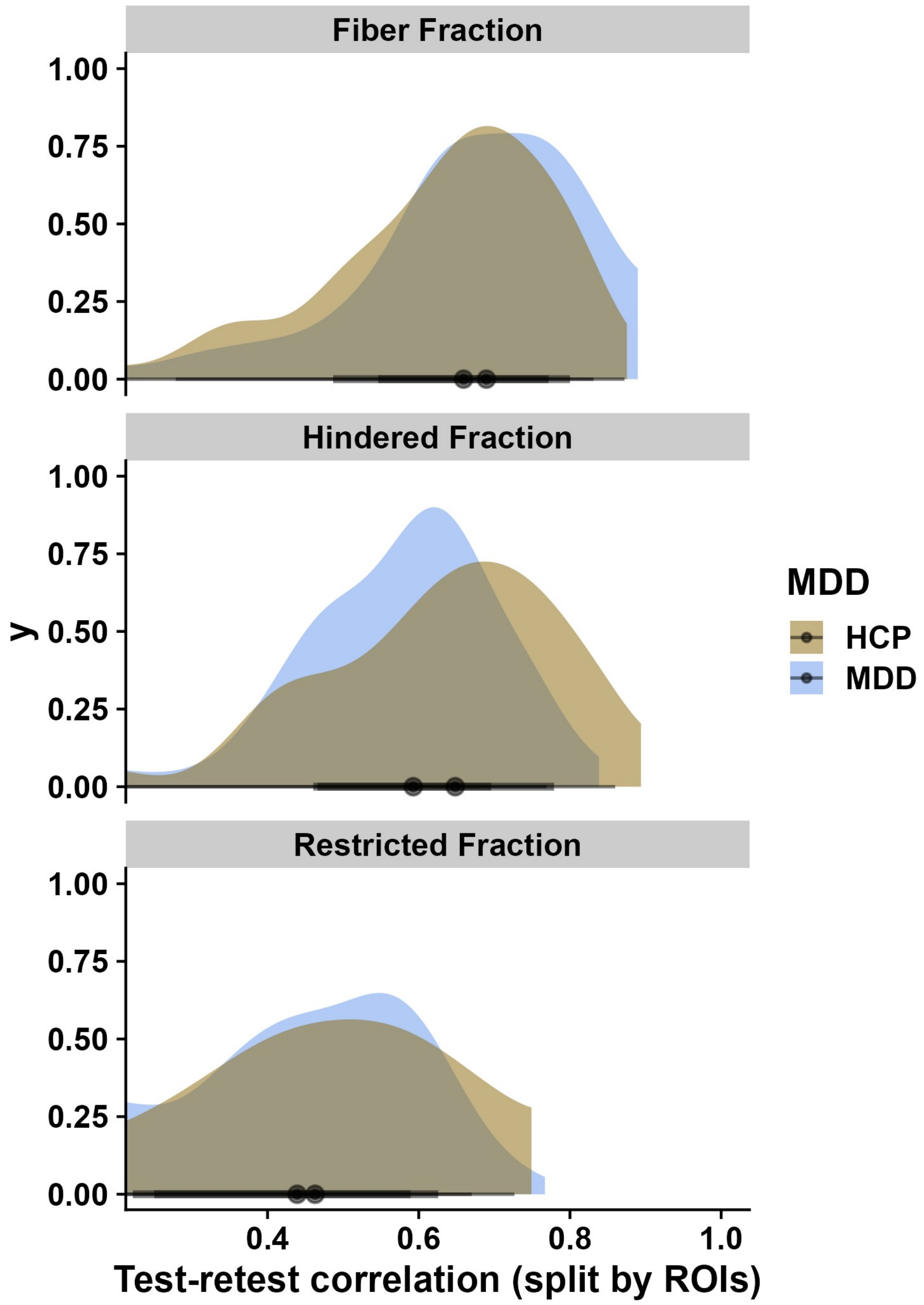


**Figure S1:** Distribution of the test-retest correlation across gray matter ROIs calculated separately for participants with major depressive disorder (MDD) vs. healthy control participants (HCP). The MDD group has lower reliability of hindered fraction across ROIs.

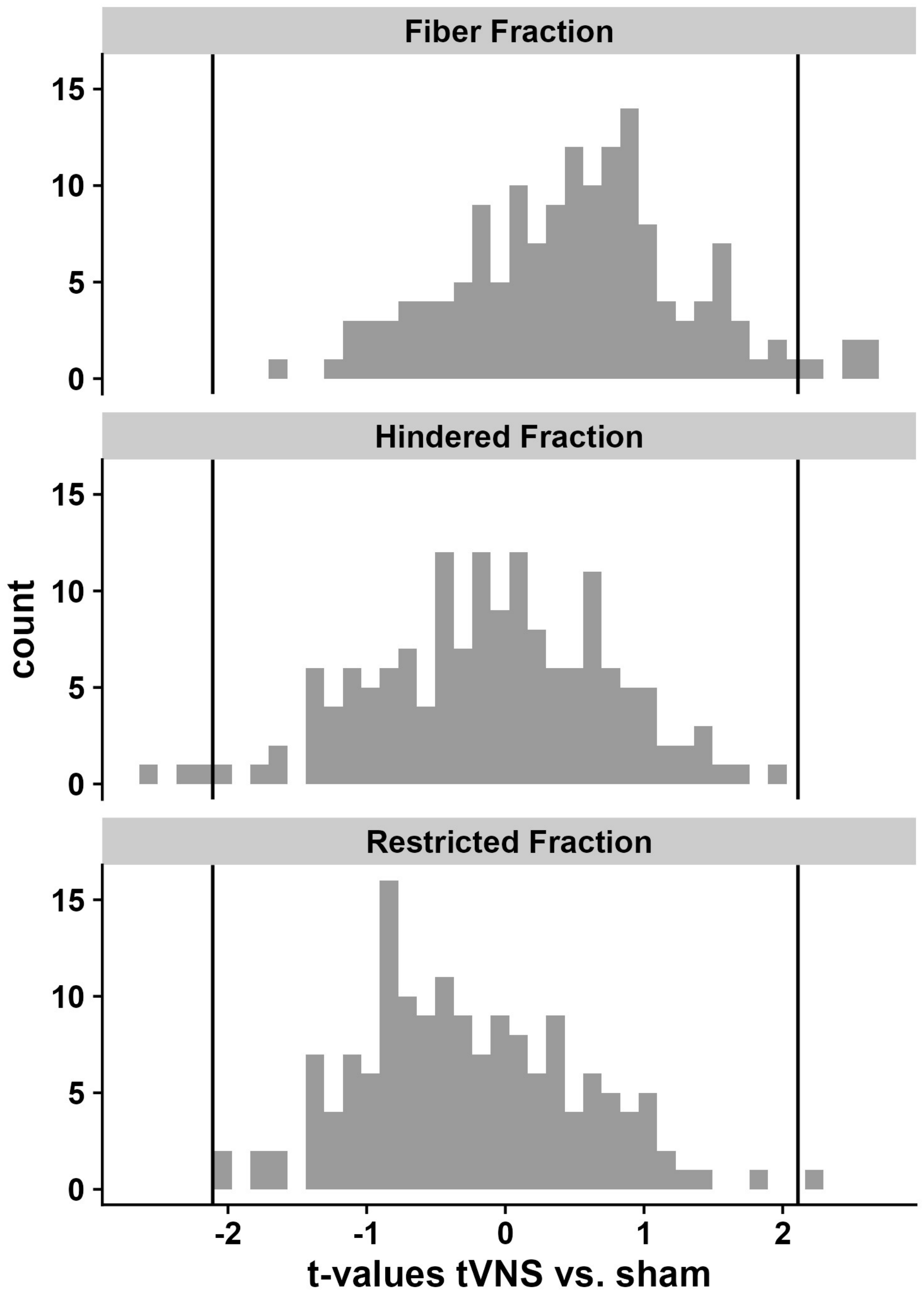


**Figure S2:** Distribution of the effect of stimulation (transcutaneous vagus nerve stimulation, tVNS vs. sham) on changes in diffusion basis spectrum imaging metrics after 1.5 hours. Distributions show t-values across gray matter regions of interest. Vertical lines indicate the critical t-value for differences that are significant with $p$<.05 on an uncorrected level. After FDR-correction, no effects remain significant ($p$s>.59).

**fMRIprep preprocessing of anatomical T1 images**

For each participant one high resolution T1 weighted (T1w) anatomical image was acquired during the first neuroimaging session. The T1w image was corrected for intensity non-uniformity (INU) with `N4BiasFieldCorrection` (Tustison et al., 2010), distributed with ANTs 2.5.0 (Avants et al., 2008, RRID:SCR_004757). The T1w-reference was then skull-stripped with a Nipype implementation of the `antsBrainExtraction.sh` workflow (from ANTs), using OASIS30ANTs as target template. The resulting brain mask was applied to the preprocessed T1-weighted image using `fslmaths` from FSL.

Volume-based spatial normalization to one standard space (MNI152NLin2009cAsym) was performed through nonlinear registration with `antsRegistration` (ANTs 2.5.0), using brain-extracted versions of both T1w reference and the T1w template. The following template was selected for spatial normalization and accessed with TemplateFlow (23.1.0, Ciric et al., 2022): ICBM 152 Nonlinear Asymmetrical template version 2009c [(Fonov et al., 2009), RRID:SCR_008796; TemplateFlow ID: MNI152NLin2009cAsym].

**Transcutaneous vagus nerve stimulation device**

Participants received either tVNS or sham according to a randomization list created prior to enrollment during both neuroimaging sessions. We used a previously established, conventional stimulation protocol (25Hz, 30s on/30s off, 250µs pulse width; tVNS R device, tVNS Technologies GmbH, Erlangen, Germany) in line with previous studies (Teckentrup et al., 2025, 2021). For tVNS, the electrode was positioned on the right ear at the cymba conchae, targeting the auricular branch of the vagus nerve (Frangos et al., 2015). Sham stimulation was applied to the earlobe, which is not innervated by the vagus nerve (Farmer et al., 2021; Peuker and Filler, 2002).

Stimulation intensity was individually adjusted for each session using a pain visual rating scale (0 = “no sensation”, 10 = “strongest sensation imaginable”). Stimulation started at 0.1mA and was increased until participants reported a stable rating of approximately 5 (“mild pricking”). This intensity was then applied throughout the session ($M_{tVNS}$=2.01±1.02mA, $M_{sham}$=2.67±1.09mA).
